# Hospital Outcome Prediction Equation (HOPE) model, version 7

**DOI:** 10.1101/2025.07.13.25330500

**Authors:** Graeme J. Duke, Steven Hirth, Adina Hamilton, Melisa Lau, Zhuoyang Li, Teresa Le, Tharanga Fernando, John D. Santamaria

## Abstract

**Background:** The hospital standardised mortality ratio (HSMR) is a simple ratio, yet, plagued by sparsity, dimensionality, over-dispersion, exclusivity, and clinical controversy. We describe the Hospital Outcome Prediction Equation, version-7 (HOPE-7) methodology, derived from jurisdictional administrative data, which addresses many of these limitations.

**Methods:** Population included all adult acute-care hospital separations in State of Victoria (population 6.8 million), Australia, over 5-years July 2018-June 2023. Multistage model development included: (a) aggregation of 12,145 principal diagnoses (reason for hospital admission) from International Classification of Disease version 10 into twenty categories ranked according to estimated risk of death; (b) fixed-effect logistic regression (with adjustment for clustering at hospital level) fitted to a randomly selected (75%) training dataset, with low-risk (case fatality rate, CFR <0.02%) diagnosis groups allocated to a zero risk category; (c) and model performance tested in (remaining 25%) validation dataset. Calibration assessed by Hosmer-Lemeshow goodness-of-fit [H_10_], Brier score, calibration plot; model discrimination by area under precision-recall (AUCPR) and receiver-operator (AUCROC) curves; and model classification at the hospital level by dispersion value [φ], standard deviation of random effect [τ]) and proportion of in-control hospitals. Ideal model characteristics: Brier score ∼0, H_10_ p-value >0.05, AUCPR>0.30, AUCROC >0.80, φ →1.0, τ →0, and <1% hospitals outside ±3SD of benchmark.

**Results:** 315 hospitals treated 12.97-million adult acute-care separations with mean CFR 0.59% (95%CI = 0.58-0.60; n=77,637). 4,211 (34.7%) principal diagnoses allocated to zero mortality-risk category (CFR 0.003%); 3,497 (28.8%) non-significant diagnoses allocated to baseline mortality risk category (CFR 0.50%); remaining 4,437 (36.5%) diagnoses aggregated into 18 ranked risk categories (CFR range: 0.08-32.5%). Final model, developed on 9.75-million records, included one continuous variable (age in years, transformed to square root); five binary demographic variables (sex, relationship status, unplanned admission, aged-care resident, hospital transfer); one interaction term (emergency-transfer); and twenty diagnosis-risk categories, of which, only one was allocated to each record. Validation cohort (n= 3.25 million) parameters included: Brier score =0.015; H_10_ =14.88 (p=0.094); AUCPR =0.28 ±0.01; AUCROC =0.90 ±0.007; φ =4.3 and τ =0.24; before and after (multiplicative) adjustment for over-dispersion 280 (88.9%) and 313 (99.1%) hospitals, respectively, remained within ±3SD benchmark over all five years.

**Conclusion:** HOPE-7, a parsimonious and pragmatic HSMR based on administrative data common to all jurisdictions, displayed satisfactory calibration, classification, and discrimination metrics, addressed many common HSMR limitations, and complements existing methods.

## INTRODUCTION

The hospital standardised mortality ratio (HSMR) is arguably the simplest and yet the most controversial of all clinical indicators[1]. It is the ratio of observed to expected deaths, where a result above unity is indicative of more deaths than predicted and, possibly, an increased patient safety risk. Death is easy to recognise, difficult to hide, and an outcome that most patients and families prefer to avoid.

The HSMR remains controversial because it has so often failed to deliver, and for valid reasons[2]. These include sparsity, diversity, dimensionality, over-dispersion, and misinterpretation. Death in hospital is uncommon (<1% even in acute care) and often (>90%) unavoidable[2–4]. Diversity of case mix and diagnoses generates dimensionality, rendering clinically robust statistical analysis problematic. The range of HSMR options (Table E1, Supplement) in Australia[5–7] and United Kingdom[8,9] and the absence of a ‘gold standard’ attest to this lack of certainty. It is no surprise that a reliable relationship between the HSMR and the quality of hospital care remains elusive[10]. It would seem prudent to abandon the HSMR as a performance metric if it were not for the equally compelling defense of its retention.

In Australia and United Kingdom most deaths occur in an acute-care hospital than any other setting[11,12]. Even though the majority are unavoidable, healthcare providers and the communities they serve are entitled to some reassurance that unexpected hospital death rates remain low - yesterday’s reassuring result provides no guarantee for tomorrow. In passing it is worth noting that most major healthcare scandals have been characterised by an increase in unexpected deaths together with the absence (and/or ignorance) of mortality monitoring. There is some evidence that continuous monitoring of mortality rates may have identified the misconduct sooner and, possibly, saved lives[13,14]. Finally, a robust HSMR also has utility for epidemiology, healthcare research, policy, and planning[15].

An ideal HSMR is inclusive of all patients, all diagnoses, and all hospitals; is parsimonious and minimises mis-specification[1,5]; and adjusts for important (patient-related) factors that influence survival but are outside the control of the healthcare service under assessment. To achieve these goals an ideal HSMR requires reliable and comprehensive data, robust risk-adjustment, adequate calibration (risk ranking), simple visualization tools, and expert clinical interpretation.

In this report we describe the methodology underlying the Hospital Outcome Prediction Equation (HOPE, version 7), an HSMR based on the above principles and suitable to any jurisdiction that collects administrative data based on the International Classification of Diseases and Health Related Problems (ICD)[16]. The focus of this report is primarily methodological rather than diagnostic or comparative.

## METHODS

### Setting

Administrative datasets in Australia are routinely extracted from medical records, encrypted by clinical coders according to national[17] and state[18] coding guidelines. Each coded record describes a unique ‘episode of care’, includes patient demographics and all (treated and documented) diagnoses, procedures, and outcomes (length of hospital stay, survival, and discharge destination). Clinical diagnoses are currently encrypted according to the Twelfth Edition of the Australian modification of ICD, version 10 (ICD-10-AM)[18].

### Population

We obtained from the Victorian Agency for Health Information[19] a copy of the jurisdictional administrative dataset for all adult (age greater than 17-years) acute-care separations from public and private sector hospitals, between July 2018 and June 2023. An encrypted ‘patient’ key permitted linkage of the index episode to identify the (hospital spell and) final outcome. We excluded records without any reported diagnosis (reason for admission), paediatric subjects (since they have a different disease profile), palliative care services (where death is an expected outcome); and rehabilitation, geriatric evaluation, and mental health (where risk profile differs). Of note, day-procedure and maternity separations were retained. Each hospital was allocated to one of six peer group categories: public tertiary referral, major metropolitan, major regional, or other; and private tertiary or other.

### Ethical considerations

This project was approved by the Department of Health Victoria and by the Eastern Health Human Research and Ethics Committee (LR19/062); the need for patient consent was waived. A TRIPOD checklist[20] is available in Table E2 (online Supplement).

### Statistical considerations

We generated purpose-written code to facilitate analysis and employed Stata/MP™ v18.0 (2023, College Station, TX) statistical software. Modelling a rare event (death) in a high dimensional dataset (over ten thousand diagnoses) in a population of over two million records per annum carries substantial risk of mis-specification, over-fit, and over-dispersion. A multiple phase approach was required to mitigate these risks: Phase-1 and −3 reduced thousands of (ICD-10-AM diagnosis codes and) candidate variables to a clinically meaningful and statistically manageable number; Phase-2 addressed the sparsity of outcome (in-hospital death); Phase-4 generated the final model; and Phase-5 assessed overall model performance.

### Phase-1: Aggregation of ICD-10-AM codes

Each ICD-10-AM principal diagnosis (primary reason for admission[15]) code was allocated to one of 406 pre-defined Clinical Diagnosis Groups (CDGs[21]). A CDG is defined as a set of ICD-10-AM diagnosis codes that describe a similar clinical condition with similar pathology, e.g. bacterial community-acquired pneumonia, cardiac failure, and chronic kidney disease. The CDG classification system is conceptually similar, but not identical, to the Clinical Classification Software (CCS) developed by the Agency for Health Research and Quality[22] which is employed in several HSMR models[8,9]. The CDG system address several inherent limitations[21] of the CCS when applied to mortality models. To aid post-hoc stratification, all admission diagnoses were allocated to their relevant CDG, although this step is redundant to the HOPE-7 methodology.

### Phase-2: Low risk separations

A substantial volume of hospital admissions, notably maternity and day-procedure admissions, are associated with a negligible case fatality rate (CFR) and their inclusion in a prediction model increases the likelihood of over-fit. To reduce this risk we identified all CDGs with an observed CFR less than 1 in 5,000 (<0.02%) and temporarily excluded them from Phase-3 and −4 but reinstated these “low-risk” separations in the final analysis and validation phase with their ‘predicted’ mortality risk set to zero.

### Phase-3: Ranking of CDG

The purpose of this phase was to identify the strength of association between each CDG and (in-hospital) death, and rank them according to their (multivariate risk-adjusted) coefficient (ß). Only those CDGs associated with a minimum of fifty cases and thirty deaths over 5-years[23] were accepted as candidate variables. A mixed-effects regression model was fitted to the outcome (in-hospital death) and adjusted for available patient demographics (age, sex, source of admission, urgency, indigenous, ethnic, and domiciliary relationship status). This (interim) model included a random intercept for hospital site to adjust for geographical differences in case-mix, services, or models of care; and a (categorical) fixed-effect covariate for each (fiscal) year to adjust for temporal changes in case-mix. Variance at the hospital level and patient level were assessed using the intraclass correlation coefficient (ICC)[24].

Following an iterative process, each candidate variable was interrogated and retained only if it (a) was a significant (p<0.157[25]) predictor of outcome and (b) improved (that is, reduced) the model’s information criteria (consistent AIC and BIC[26]); or (c) strengthened the coefficient of a more dominant covariate. Of note, we did not employ automated (forward or backward) step-wise selection methods[5,6] which may inadvertently discard important covariates[25,27].

Statistically significant CDGs were ranked and grouped and by risk coefficient (ß) using the Stata command “*xtile*”, with the final number of categories (and the ß range for each) determined by the effect on the model’s information criteria. It is important to note that the coefficients generated from this interim (Phase-3) model were not employed in the final (Phase-4) model.

### Phase-4: Final model

The HOPE-7 model employed a generalised linear model with a logit link function fitted to in-hospital death (outcome) while the candidate variables were restricted to significant patient demographics and the ranked diagnosis groups from Phase-3. The final HOPE-7 model (differed from the interim Phase-3 model and) excluded variables pertaining to hospital site and year of admission. The form of the estimator was,

> *logit death age male aged-care emergency transfer single interaction rank-1 rank-2 … rank-i, vce(cluster hospital)*

where *logit* represents the Stata command for a standard logistic regression estimator fitted to the outcome; *death* is the binary outcome of in-hospital death; *age* in years transformed to the square-root, *male* sex, *aged-care* resident, *emergency* admission status, hospital *transfer* to higher level of care, *single* (relationship) status, and *interaction* term for unplanned inter-hospital transfer; *rank-1* to r*ank-i* represent the ranked categories of CDGs identified in Phase-3; and standard errors adjusted for clustering at the *hospital* level.

### Phase-5: Model validation

Validation of the HOPE-7 model included discrimination, calibration, classification, and dispersion metrics[27]. A random 75:25 split into training and validation cohorts permitted coefficients to be generated from the former and applied to the latter for assessment of model fit. The Brier score and Hosmer-Lemeshow goodness-of-fit statistic (with equal sized deciles [H_10_]) for each fiscal quarter were employed to assess calibration[27]. The user-written command *calibrationbelt*[28] furnished a visual and statistical assessment of calibration. Discrimination was reported as the area under the receiver operator characteristic curve (AUCROC) and, given the unbalanced dataset (that is, survivors >> deaths), the precision recall curve (AUCPRC) was assessed. An ideal model will produce a Brier score approximating zero, an H_10_ associated p-value >0.05, and AUCROC >0.80 and AUCPRC >0.30.

Since the primary purpose of a HSMR is assessment of patient outcome at the provider level[1,5], we assessed hospital classification using HOPE-7 and dispersion characteristics in the following manner. The final model was recalibrated to each fiscal year - akin to generating an annual benchmark - and the standardised mortality ratio (SMR) for each hospital in each fiscal quarter was compared to the benchmark (with control limit precision determined by the number of predicted deaths) employing the user-written command *funnelinst*[29]). Over-dispersion was quantified by the dispersion value (φ) and the standard deviation of the random effect (τ)[30]. Re-classification was assessed by comparing the number of hospitals with a CFR outside, and a SMR inside, ±3SD[27].

Applying an ideal prediction model to a group of ideal hospitals with a uniform high standard of care we would expect φ to approximate unity, τ to approximate zero, and fewer than 1% of hospital SMR values to exceed ±3SD of the benchmark. Sensitivity analyses of calibration, discrimination, and dispersion were undertaken for each hospital peer group and each fiscal year.

## RESULTS

In 2023 the estimated population in the State of Victoria was 6.81 million (with 5.28 million adults over 17-years of age [11]) served by over 350 health services. Over the five-year study period, 315 (public and private sector) acute-health services treated 3.19 million adults and 12.97 million separations; with 179 (56.8%) hospitals reporting 77,637 deaths for a grand mean CFR of 0.59 (95%CI = 0.58-0.60; hospital CFR range: 0.003 to 18.2) deaths per 100 separations (Figure E1, Supplement).

From the 14,470 available ICD-10-AM diagnosis codes in the Twelfth Edition[18], a total of 12,145 (83.9%) were employed as principal diagnoses by clinical coders over the 5-years. These codes were allocated to one of pre-defined 406 CDG sets[21] (candidate variables). After exclusion of 45 (11%) low-risk (Phase-2) and 198 non-significant (Phase-3) CDGs there were 163 CDGs identified as significant predictors of outcome, which were further aggregated into nineteen ranked categories in Phase-3. For example: Rank-1 includes any CDG with ß < −0.9; Rank-2 includes ß range of −0.9 to −0.5; Rank-3 ß = −0.5 - 0; Rank-4 ß = 0 for; Rank-5 ß = 0 - 0.4; Rank-6 ß = 0.4 - 0.7; and so forth, to Rank-18 ß = 4.1-4.4; and Rank-19 ß >4.4. All low-risk CDGs were allocated zero risk (Rank-0). Thus, Phase-1 and −3 condensed over twelve thousand (ICD-10-AM) diagnoses into twenty risk-categories.

For the final model (Phase-4) each record was assigned eight covariates: six demographic variables (age, sex, unplanned admission, aged-care resident, transfer source, single/relationship status), one interaction term (unplanned inter-hospital transfer) and one of the twenty ranked risk-categories. Tables E3 and E4 (Supplement) provide further detail and worked examples, respectively.

For the validation phase, the study population was randomly divided into training (n=9.75 million) and validation (n=3.25 million) cohorts. The calibration plot for the validation cohort is displayed in Figure 2. Over the period of (twenty) fiscal-quarters in the validation dataset, the mean Brier score was 0.015 (standard deviation [SD] 0.002) and the mean H_10_ statistic was 14.88 (p=0.094; Table E4, Supplement). The AUCROC was 0.895 (95%CI = 0.874-0.915), AUCPRC was 0.277 (95%CI =0.247-0.306); and the values for each peer group are presented in Table 2 and Figure 3. Sensitivity analyses for each fiscal year (Figure E2) and peer group (Figure E3) are available in the online Supplement.

**Figure 1.**
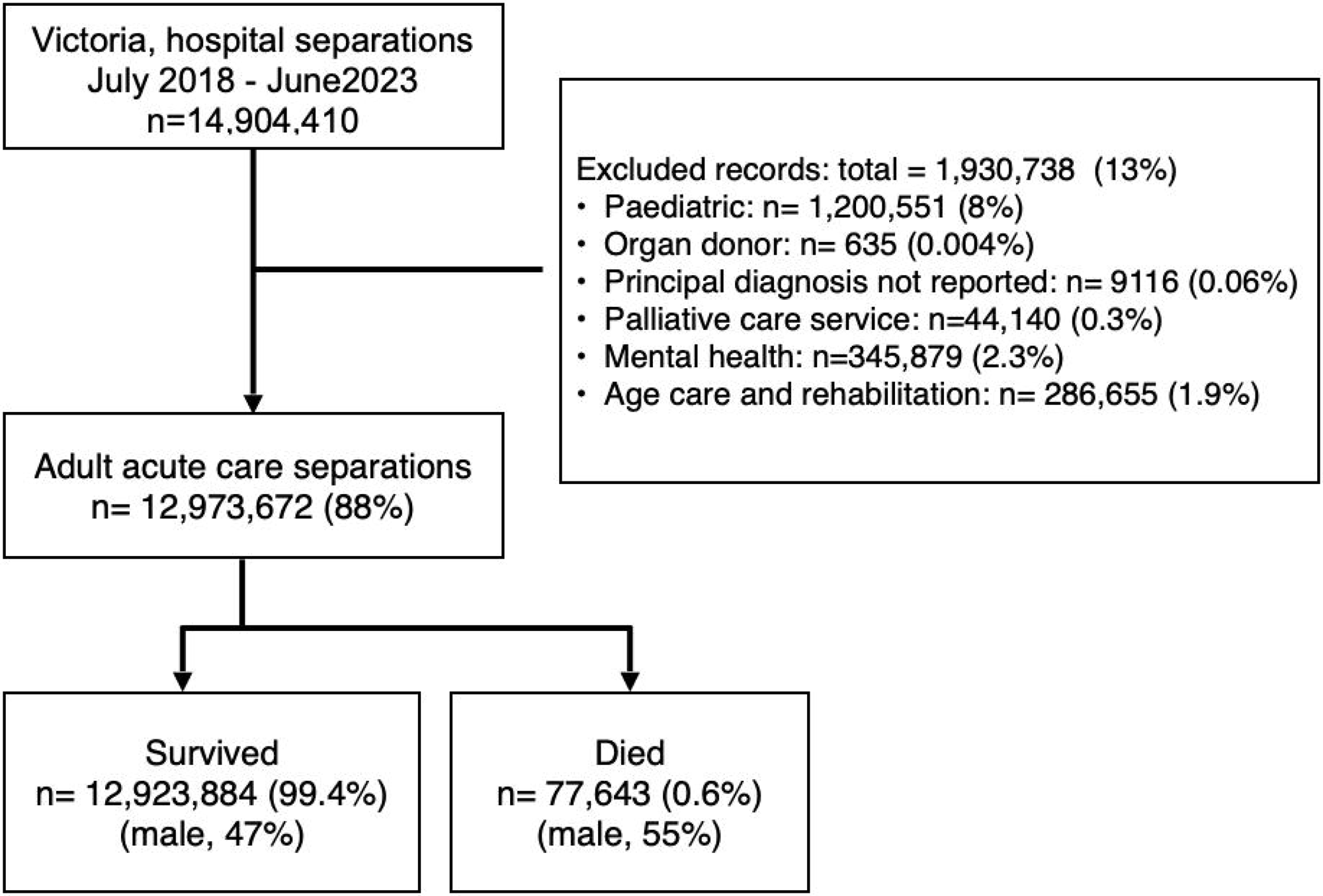
STROBE diagram of study population. Note: more than one reason for exclusion possible.

**Figure 2.**
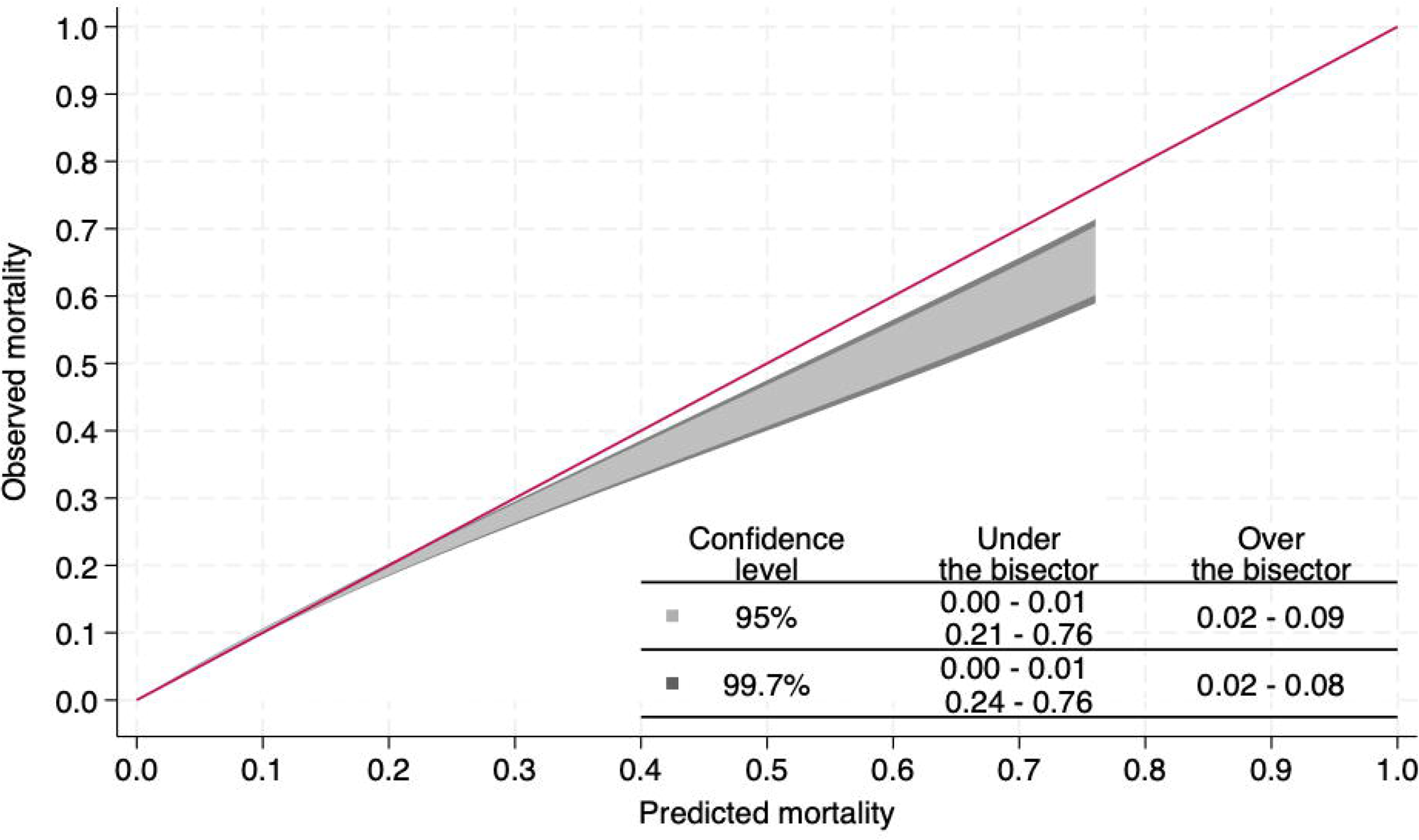
HOPE-7 calibration plot in external validation dataset (n=1.09 million), excluding low-risk admissions. Test statistic= 99.4 (p<0.001). Bisector (red line) identifies ideal model.

**Figure 3.**
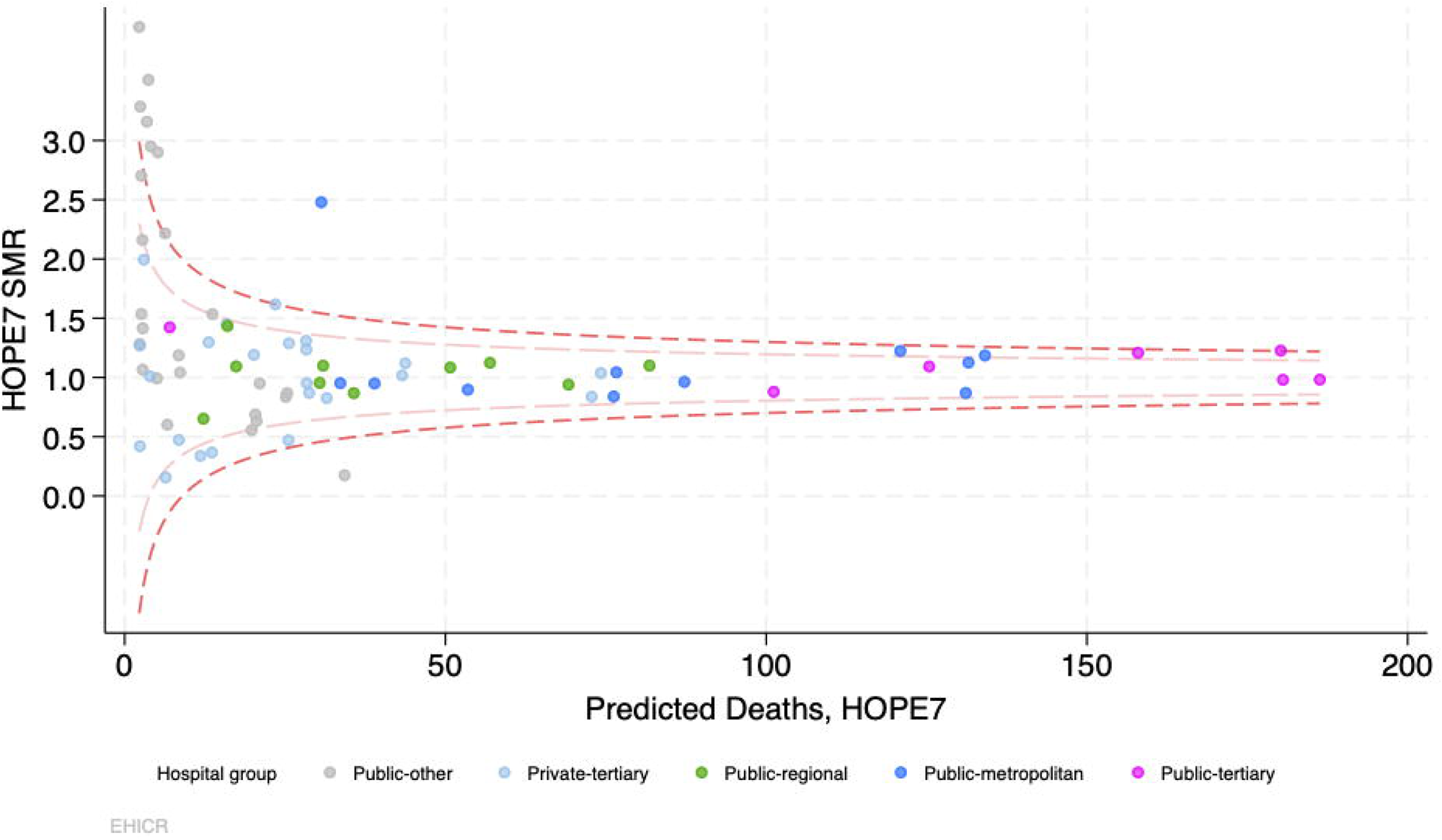
Funnel plot of HOPE-7 HSMR in validation dataset for fiscal year ending 30th June 2023 compared to the state benchmark (dashed lines represent ±2SD and ±3SD) without adjustment for over-dispersion (□ = 4.0 and □ =0.29); n=543,121 separations, 3,374 deaths; 278 hospitals (solid circles); hospitals with ≤2 deaths not shown.

To test model classification, data for 5,515 ‘hospital-quarters’ over 5-years was available for anlysis after excluding 785 (12%) hospital-quarters from 142 separate hospitals reporting no deaths. During this time period, 280 (88.9%) hospitals and 5,365 (97.3%) ‘hospital-quarters’ remained within ±3SD of the relevant benchmark. The dispersion characteristics, □ = 4.3 and □ = 0.24 (Table E5, Supplement) indicated the presence of over-dispersion. Following multiplicative[28] adjustment, 313 (99.1%) hospitals were within ±3SD, and 305 (96.5%) within ±2SD, of the state benchmark. Forty hospitals with an unadjusted CFR outside ±3SD were reclassified as inliers after risk-adjustment; but none of the CFR inliers were reclassified as SMR outliers. The (unconditional) ICC values at the hospital and patient level were 0.05 (95% CI, 0.03-0.10) and 0.61 (95% CI, 0.60-0.62), respectively.

## DISCUSSION

### Principal findings

This report describes the methodology and performance characteristics of the Hospital Outcome Prediction Equation model, version seven (HOPE-7): a parsimonious and inclusive HSMR that displays acceptable discrimination, calibration, and classification metrics (Table-2, Figures E2-E4) for the purpose of monitoring hospital performance and patient safety.

Like other HSMR methods HOPE-7 is derived from administrative data and ICD-10 diagnosis codes and therefore widely applicable. It has several features that render it complementary to existing methods. HOPE-7 is inclusive of all acute-care (public and private) hospitals, all adult acute-care separations both low-risk (day-procedure and maternity) and high-risk (palliative care patients managed in acute-care beds) and has few exclusions. While the methodology appears complex, the HOPE-7 model is not. It requires only eight covariates and one formula, includes all principal diagnoses rather than requiring separate prediction models for each[8,9].

### Strengths and limitations

HOPE-7 is parsimonious and applicable to any jurisdiction where administrative data are based on ICD-10, and applicable to hospitals of different size, activity, case mix, and peer group. The ranking of diagnoses is based on clinically meaningful diagnosis groups (CDG[21]) rather than the original ICD-10-AM code[18]. By design, the model excludes (a-priori) hospital-related factors under assessment (treatment-dependent variables) such as clinical interventions and therapeutic choices, complications, or length of stay[6].

Our methodology has several limitations. Administrative data based on ICD codes lack granularity (clinical severity) and are dependent on the accuracy of coding and, more importantly, the quality of clinical documentation from which they were derived[18]. Although the multi-phase methodology appears somewhat cumbersome this is common to most HSMR models and, arguably, unavoidable. Diversity of hospital case-mix, the dimensionality of clinical data (thousands of diagnosis codes[21]), and the limitations of administrative data extracted from clinical documentation of the variable quality, all conspire to obfuscate analysis. They are not designed to guide clinical decisions (e.g. low AUCPR values, Table-2.)

### Interpretation

Although the primary purpose of this report is methodological, the clinical implications warrant brief comment. The results suggest that there was a high degree of uniformity in patient outcomes under the current model of care in the state of Victoria, despite the diversity in patient acuity and providers. Of note, a substantial period of this investigation straddled the recent SARS-CoV2 pandemic (Table 1) during which profound temporary changes in model of care and hospital case-mix arose.

**Table 1.**
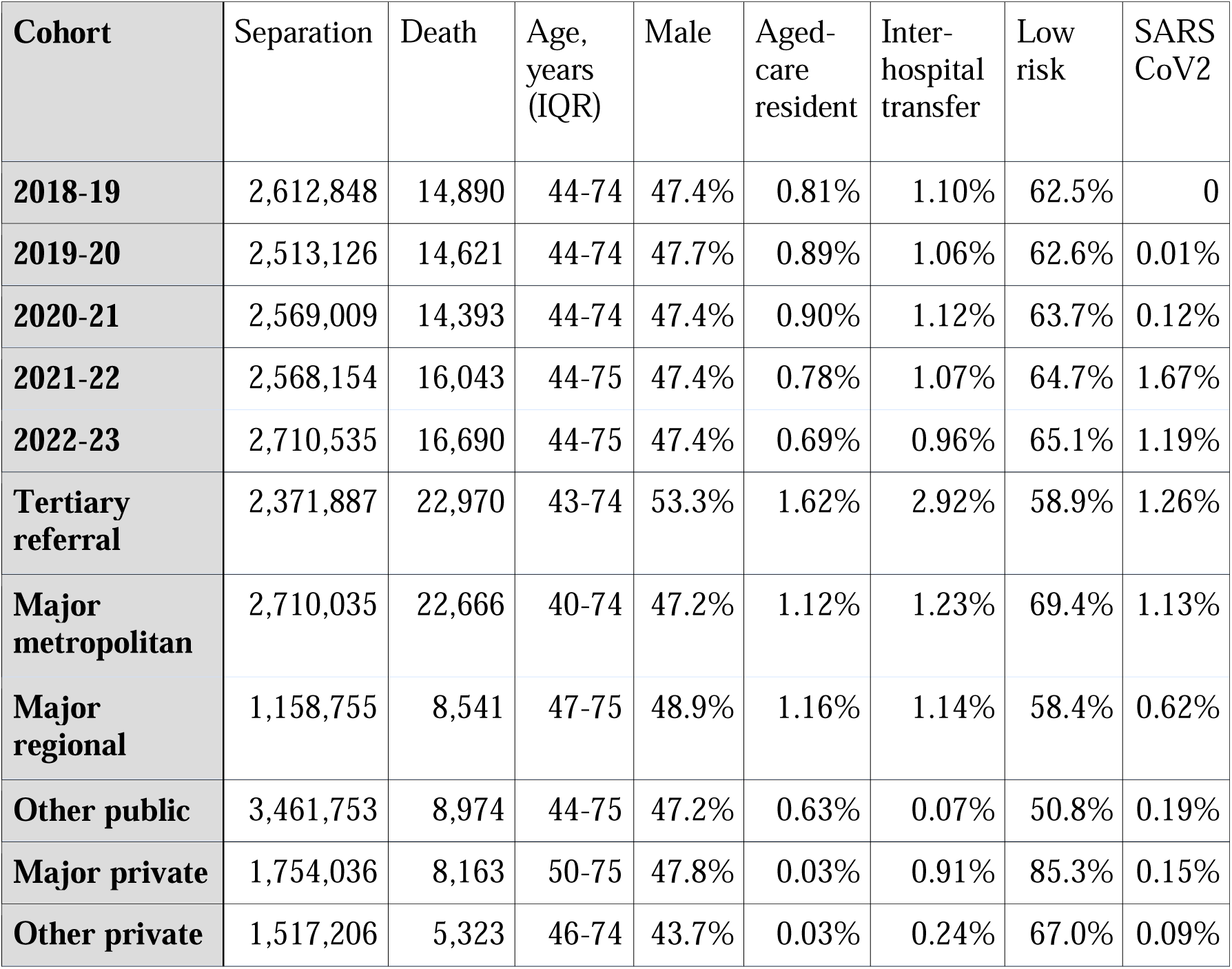
Population demographics according to fiscal year and hospital peer groups. IQR = interquartile range; Transfer = proportion transferred from similar or lower service level; Low risk = proportion of separations allocated to Clinical Diagnosis Group with CFR <0.02%; SARS CoV2 = pandemic infection with severe acute respiratory syndrome coronavirus-2.

**Table 2.** Outcomes and HOPE-7 model performance in validation dataset. aLOS = average length of hospital stay; CFR = case fatality rate; AUCPRC = area under receiver precision recall curve; AUCROC = area under receiver operator characteristic curve; H_10_ = Hosmer-Lemeshow chi-squared statistic with equal size deciles.

| Patient Group | aLOS,<br>days | CFR | AUCPRC | AUCROC | Brier<br>score | H<sub>10</sub> |
| --- | --- | --- | --- | --- | --- | --- |
| 2018-19 | 1.593 | 0.57% | 0.273 | 0.899 | 0.014 | 21.6 |
| 2019-20 | 1.592 | 0.58% | 0.278 | 0.894 | 0.014 | 27.0 |
| 2020-21 | 1.568 | 0.56% | 0.264 | 0.899 | 0.014 | 18.4 |
| 2021-22 | 1.550 | 0.62% | 0.290 | 0.896 | 0.016 | 34.6 |
| 2022-23 | 1.591 | 0.62% | 0.281 | 0.888 | 0.016 | 42.6 |
| Tertiary referral | 2.074 | 0.97% | 0.305 | 0.889 | 0.017 | 18.0 |
| Major metropolitan | 1.827 | 0.84% | 0.344 | 0.900 | 0.015 | 29.9 |
| Major private | 2.091 | 0.47% | 0.263 | 0.892 | 0.011 | 71.9 |
| Other private<br>hospital | 1.469 | 0.35% | 0.228 | 0.924 | 0.010 | 50.0 |
| Major regional | 1.868 | 0.74% | 0.234 | 0.878 | 0.017 | 49.2 |
| Other public hospital | 0.737 | 0.26% | 0.233 | 0.883 | 0.017 | 176.8 |

Moreover, hospital survival is predominantly determined by patient factors (ICC, 0.61) rather than hospital factors (ICC, 0.05) despite the wide range in crude hospital CFR (Figure E1). This is consistent with published clinical reviews of hospital deaths[2–4]. This conclusion does not, however, negate the importance of continuous monitoring of hospital mortality and unexpected deaths.

### Implications for policy, practice and research

The HOPE-7 model complements, rather than supercedes, other mortality and outcome-based monitoring tools by addressing common limitations. All mortality prediction models employed in Australia[5–7] and United Kingdom[8,9] have a purpose, despite their limitations (Table E1, Supplement). A comprehensive review of current HSMR options is beyond the scope of this report. These methods function as screening rather than diagnostic tools[1,8]. Such models seek to identify ‘signals of interest’ that may warrant further attention, where pejorative terms such as “outlier” are best avoided. They generate probabilities based on groups not individuals - they are formative rather than summative, descriptive rather than prescriptive. Access to more than one HSMR method facilitates the ‘triangulation’ of signals of interest or concern.

### Conclusions

The HOPE-7 is a parsimonious hospital mortality prediction model derived from administrative data which provides a pragmatic method for epidemiology and monitoring mortality in adult acute health services and seeks to address common pitfalls that continue to pursue the HSMR.

## Data Availability

All data produced in the present work are contained in the manuscript. Source data are openly available from Victorian Agency for Health Information at https://vahi.freshdesk.com/support/home

## REFERENCES

1. Scott IA, Brand CA, Phelps GE, Barker AL, Cameron PA. Using hospital standardised mortality ratios to assess quality of care — proceed with extreme caution. Med J Australia 2011;194(12):645–8.

2. Rodwin BA, Bilan VP, Merchant NB, et al. Rate of Preventable Mortality in Hospitalized Patients: a Systematic Review and Meta-analysis. J Gen Intern Med 2020;35(7):2099–106.

3. Kobewka DM, Walraven C van, Taljaard M, Ronksley P, Forster AJ. The prevalence of potentially preventable deaths in an acute care hospital. Medicine 2017;96(8):e6162.

4. Hogan H, Zipfel R, Neuburger J, Hutchings A, Darzi A, Black N. Avoidability of hospital deaths and association with hospital-wide mortality ratios: retrospective case record review and regression analysis. Br Méd J 2015;351:h3239.

5. Niven P, Good N, Khanna S, Jayasena R. Hospital Standardised Mortality Ratio (HSMR) for summary mortality reporting. Useability and alternative approaches. Report prepared for Victorian Agency for Health Information, 2021. Commonwealth Scientific and Industrial Research Organisation (CSIRO). Available from. Accessed December 2024.

6. National core, hospital-based outcome indicator specification, 2019. Version 3. Available from https://www.safetyandquality.gov.au/sites/default/files/migrated/Specification-for-National-core-hospital-based-outcome-indicator-CHBOI-V3-April-2019.pdf. Accessed December 2024.

7. Cattell R. AP-MortR v1.1. 2018. [Personal communication] Available upon application from: https://www.healthroundtable.org/Join-Us/About-Us/Contact-Us.

8. Summary Hospital-level Mortality Indicator: Indicator specification. NHS December 2024. Available at https://digital.nhs.uk/data-and-information/publications/statistical/shmi/2024-12. Accessed December 2024

9. Scottish Hospital Standardised Mortality Ratio (HSMR). Methodlogy and Specification Document, 2016. Public Health Scotland. Available at www.publichealthscotland.scot/media/24426/hsmr_2016_technical_specification.pdf. Accessed December 2024.

10. Pitches DW et al. What is the empirical evidence that hospitals with higher-risk adjusted mortality rates provide poorer quality care? A systematic review of the literature. BMC Health Serv Res 2007;7(1):91.

11. Australian Bureau of Statistics. Classifying Place of Death in Australian Mortality Statistics. Canberra: ABS; 2021 April 14. Available from: https://www.abs.gov.au/statistics/research/classifying-place-death-australian-mortality-statistics. Accessed December 2024.

12. Classification of place of death. A technical bulletin from the National End of Life Care Intelligence Network. Public Health England. Available at https://assets.publishing.service.gov.uk/media/5d7f9e0440f0b61c7a6640d4/Classification_Place_Death_report.pdf. Accessed December 2024

13. Spiegelhalter D et al. Risk-adjusted sequential probability ratio tests: applications to Bristol, Shipman and adult cardiac surgery. International journal for quality in health care. 2003;15(1):7–13.

14. Pilcher DV, Hoffman T, Thomas C, Ernest D, Hart GK. Risk-adjusted continuous outcome monitoring with an EWMA chart: could it have detected excess mortality among intensive care patients at Bundaberg Base Hospital. Critical care and resuscitation 2010;12(1):36–41.

15. Jarman B, Pieter D, Veen AA van der, et al. The hospital standardised mortality ratio: a powerful tool for Dutch hospitals to assess their quality of care? Qual Saf Heal Care 2010;19(1):9.

16. Coding Rules and FAQs for ICD-10-AM/ACHI/ACS Twelfth Edition. Independent Health and Aged Care Pricing Authority 2023. Available at https://www.ihacpa.gov.au/. Accessed December 2024.

17. Victorian Admitted Episodes Dataset (VAED) manual 2022-2023. Department of Health, Victoria. June 2022 Available from: https://www.health.vic.gov.au/data-reporting/victorian-admitted-episodes-dataset. Accessed December 2024.

18. ICD-10-AM/ACHI/ACS Twelfth Edition. Independent Health and Aged Care Pricing Authority. September 2023. Available at https://www.ihacpa.gov.au/resources/icd-10-amachiacs-twelfth-edition. Accessed December 2024.

19. Victorian Agency for Health Information. https://vahi.freshdesk.com/support/home. Accessed December 2024.

20. Collins GS, Moons KGM, Dhiman P, et al. TRIPOD+AI statement: updated guidance for reporting clinical prediction models that use regression or machine learning methods. BMJ 2024;385:e078378.

21. Duke GJ, Hirth S, Santamaria JD, et al. Clinically meaningful categorisation of ICD-10-AM (Australian modification). Health Inf Manag J 2024; doi: 10.1177/18333583241296224.

22. Clinical Classifications Software Refined, AHRQ. Available at https://hcup-us.ahrq.gov/toolssoftware/ccsr/ccs_refined.jsp. Accessed December 2024.

23. Peduzzi P, Concato J, Kemper E, Holford TR, Feinstein AR. A simulation study of the number of events per variable in logistic regression analysis. J Clin Epidemiology 1996;49(12):1373–9.

24. Liljequist D, Elfving B, Skavberg Roaldsen K. Intraclass correlation: a discussion and demonstration of basic features. PLoS ONE 2019; 14: e0219854.

25. Heinze G, Dunkler D. Five myths about variable selection. Transpl Int 2017; 30: 6–10.

26. Kuha J. AIC and BIC: comparisons of assumptions and performance. Sociol Methods Res 2016; 33: 188–229.

27. Steyerberg EW, Vickers AJ, Cook NR, et al. Assessing the Performance of Prediction Models. Epidemiology 2010;21(1):128–38.

28. Nattino G, Lemeshow S, Phillips G, Finazzi S, Bertolini G. Assessing the Calibration of Dichotomous Outcome Models with the Calibration Belt. Stata J 2018;17(4):1003–14.

29. Linden A, Spiegelhalter DJ. Funnelinst: Stata module for generating a funnel plot to compare institutional performance, 2024. Available at https://ideas.repec.org/c/boc/bocode/s459283.html. Accessed December 2024.

30. Spiegelhalter DJ. Handling over-dispersion of performance indicators. Qual Saf Heal Care 2005;14(5):347.

