## Supplement for "Hospital Outcome Prediction Equation (HOPE) model, version 7"


**Table E1** Comparison of HSMR models available in Australia, p2.

**Table E2.** TRIPOD statement for reporting of prediction models and algorithms, p3-5.

**Figure E1.** Scatter plot of hospital case fatality rates, 2019-2023, p6.

**Table E3.** HOPE-7 model variables, p7-9.

**Table E4.** Example of HOPE-7 calculation, p10.

**Figure E2.** Calibration plots across hospital peer groups, validation dataset, p11.

**Figure E3.** Calibration plots across fiscal years, validation dataset, p12.

**Figure E4.** Receiver-operator characteristic plots for HOPE-7 across hospital peer groups, p13.

**Table E5.** Activity, calibration and dispersion metrics for each fiscal quarter, p14.

**Table E1** Comparison of major HSMR models available in Australia and United Kingdom.

| HSMR method: | Ideal | CHBOI | APMort-R | SHMI | HSMR (Scotland) | HOPE-7 |
|---|---|---|---|---|---|---|
| Country of origin | | Australia | Australia | England, Wales | Scotland | Australia |
| Hospital sector | All | Public | Sub-scribers | Public | Sub-scribers | Public & private |
| Day procedures included | Yes | No | No | No | No | Yes |
| Maternity included | Yes | No | Yes | No | No | Yes |
| Hospice (care type 8) | No | No | No | No | No | No |
| Palliative admission to acute care | No | Yes | Yes | Yes | Yes | Yes |
| Other care types included | | GEM | | | | |
| Age range | ≥18 years | >29-days | ≥18 | All | | ≥18 |
| Outcome interval | Flexible | Acute episode | Acute episode | Hospital spell | 30-day | Hospital stay |
| Diagnoses | All | Top 80% | All | All | All | Principal diagnoses |
| ICD-10 classification | Clinical | ICD-10-AM | ICD10-AM | CCS, AHRQ | CCS, AHRQ | CDG |
| Model | Simple | Complex | Complex | Complex | | Simple |
| Single or multiple models | Single | Single | Single | Multiple, n= 144 | | Single |

HSMR= hospital standardized mortality rate; CHBOI = National core, hospital-based outcome indicator[6]; APMort-R= Admitted Patient Mortality Risk, Health Roundtable[7]; SHMI = Summary Hospital-level Mortality Indicator[8]; GEM= geriatric evaluation and management; CCS AHRQ = Clinical Classification Software, Agency for Health Research and Quality[22]; ICD10 = International Classification of Disease and Health-Related Problems, version ten[18]; CDG=Clinical Diagnosis Groups[21]; LOS = length of hospital stay.

| | | TRIPOD Checklist Item[21] | Page |
|---|---|---|---|
| **Title and abstract** | | | |
| Title | 1 | Identify the study as developing and/or validating a multivariable prediction model, the target population, and the outcome to be predicted. | Title |
| Abstract | 2 | Provide a summary of objectives, study design, setting, participants, sample size, predictors, outcome, statistical analysis, results, and conclusions. | Abstract |
| **Introduction** | | | |
| Background and objectives | 3a | Explain the medical context (including whether diagnostic or prognostic) and rationale for developing or validating the multivariable prediction model, including references to existing models. | Introduction |
| | 3b | Specify the objectives, including whether the study describes the development or validation of the model or both. | Introduction |
| **Methods** | | | |
| Source of data | 4a | Describe the study design or source of data (e.g., randomized trial, cohort, or registry data), separately for the development and validation data sets, if applicable. | Methods |
| | 4b | Specify the key study dates, including start of accrual; end of accrual; and, if applicable, end of follow-up. | Methods |
| Participants | 5a | Specify key elements of the study setting (e.g., primary care, secondary care, general population) including number and location of centres. | Methods |
| | 5b | Describe eligibility criteria for participants. | Methods |
| | 5c | Give details of treatments received, if relevant. | Methods |
| Outcome | 6a | Clearly define the outcome that is predicted by the prediction model, including how and when assessed. | Methods, Reuslts |
| | 6b | Report any actions to blind assessment of the outcome to be predicted. | n/a |
| Predictors | 7a | Clearly define all predictors used in developing or validating the multivariable prediction model, including how and when they were measured. | Results |
| | 7b | Report any actions to blind assessment of predictors for the outcome and other predictors. | Methods |
| Sample size | 8 | Explain how the study size was arrived at. | Methods |

| | | | |
|---|---|---|---|
| Missing data | 9 | Describe how missing data were handled (e.g., complete-case analysis, single imputation, multiple imputation) with details of any imputation method. | Methods |
| Statistical analysis methods | 10a | Describe how predictors were handled in the analyses. | Methods |
| | 10b | Specify type of model, all model-building procedures (including any predictor selection), and method for internal validation. | Methods |
| | 10d | Specify all measures used to assess model performance and, if relevant, to compare multiple models. | Methods |
| Risk groups | 11 | Provide details on how risk groups were created, if done. | Methods |
| **Results** | | | |
| Participants | 13a | Describe the flow of participants through the study, including the number of participants with and without the outcome and, if applicable, a summary of the follow-up time. A diagram may be helpful. | Fig 1 |
| | 13b | Describe the characteristics of the participants (basic demographics, clinical features, available predictors), including the number of participants with missing data for predictors and outcome. | Table 1 |
| Model development | 14a | Specify the number of participants and outcome events in each analysis. | Results |
| | 14b | If done, report the unadjusted association between each candidate predictor and outcome. | - |
| Model specification | 15a | Present the full prediction model to allow predictions for individuals (i.e., all regression coefficients, and model intercept or baseline survival at a given time point). | Appendix 3 |
| | 15b | Explain how to the use the prediction model. | Table E5 |
| Model performance | 16 | Report performance measures (with CIs) for the prediction model. | Table E4 |
| **Discussion** | | | |
| Limitations | 18 | Discuss any limitations of the study (such as nonrepresentative sample, few events per predictor, missing data). | Discussion |
| Interpretation | 19b | Give an overall interpretation of the results, considering objectives, limitations, and results from similar studies, and other relevant evidence. | Discussion |
| Implications | 20 | Discuss the potential clinical use of the model and implications for future research. | Discussion |

| Other information | | | |
|---|---|---|---|
| Supplementary information | 21 | Provide information about the availability of supplementary resources, such as study protocol, Web calculator, and data sets. | Supplement |
| Funding | 22 | Give the source of funding and the role of the funders for the present study. | Nil |

**Table E2** TRIPOD[20] statement for reporting of prediction models and algorithms.

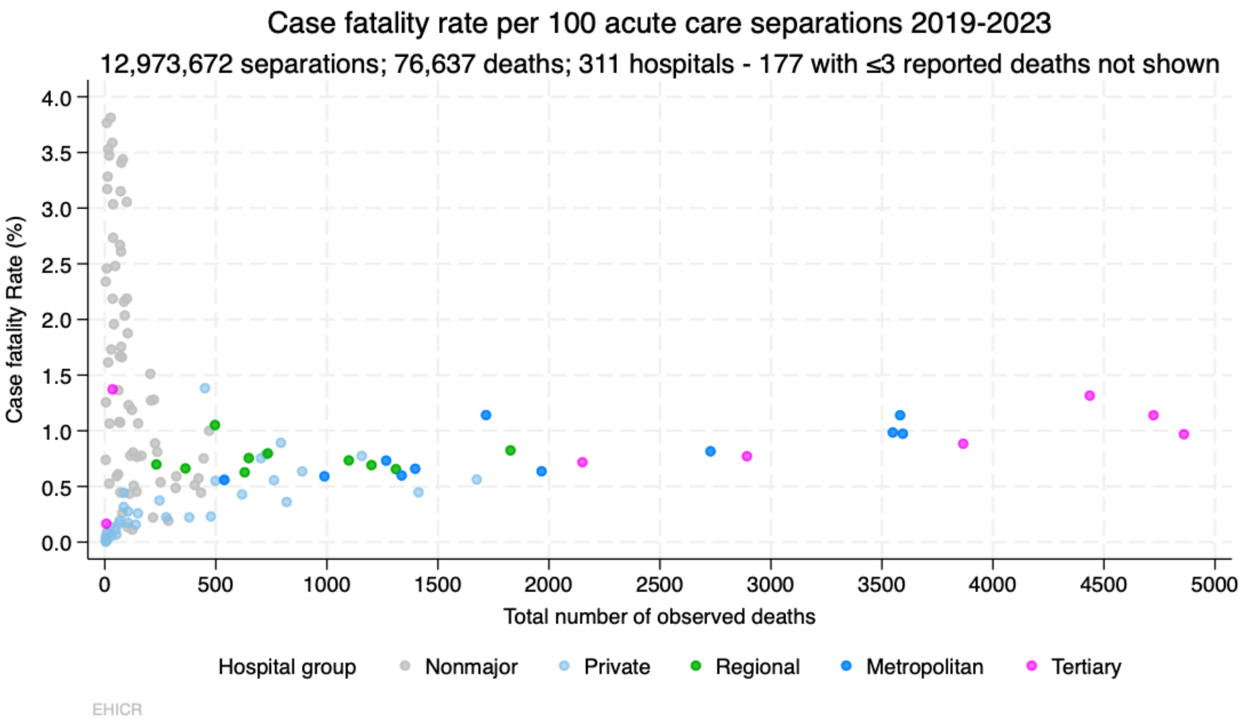


**Figure E1**

Scatter plot of hospital case fatality rates for acute-care separations over the 5-years, July 2018 - June 2023. Hospitals reporting fewer than fifty separations or three deaths, not shown.

| Variable | ß coefficient (95%CI) | ICD10-AM, n | Comment |
|---|---|---|---|
| Age (years) | 0.730, 0.854 | | Square root transformation |
| Sex | 0.087, 0.123 | | male=1, female=0 |
| Single | 0.144, 0.250 | | Partner, spouse = 0 |
| Unplanned | 0.553, 0.888 | | Admission type =emergency |
| Aged-care | 0.421, 0.597 | | Aged-care resident prior to admission |
| Up-transfer | 0.599, 1.105 | | Interhospital transfer from similar or lower level of care |
| Transfer to ED | -0.772, -0.193 | | Interaction term: transfer + unplanned admission |
| **Diagnosis rank** | | **ICD10-AM, n** | **Examples of admission diagnoses** |
| (Rank-0) | (zero risk) | 4,211 | Maternity, elective day-case procedures, ambulatory dialysis, chemotherapy, radiotherapy, all CDG with CFR<0.02% |
| Rank-1 | -2.044, -1.700 | 1,070 | Chest pain for investigation, osteoarthritis (joint replacement). |
| Rank-2 | -0.779, -0.605 | 645 | Syncope or cranial nerve disorder; hernia or prolapse without obstruction, degenerative spinal, facial trauma, hydronephrosis, psoriasis, inflammatory arthritis; urinary retention. |
| Rank-3 | -0.395, -0.194 | 158 | Nausea, vomiting, abdominal pain; asthma, upper respiratory infection, limb trauma, open wound, minor traumatic brain injury |
| Rank-4 | 0 | 3,497 | (All non-significant CDG) |
| Rank-5 | 0.284, 0.430 | 136 | Fever/pyrexia, acquired anameia, cellulitis, trauma after fall, cholelithiasis, post-operative wound, prosthesis complication, (non-melanotic) skin cancer |
| Rank-6 | 0.561, 0.699 | 321 | Hypotension, bradycardia, hyperkalaemia, or hypoglycaemia; inflammatory colitis/mucositis, infectious gastroenteritis, autoimmune disorder, epilepsy, maxillofacial disorder, lumbar spine injury, pericarditis, pyelonephritis, post-operative complication, viral infections. |

| Variable | ß coefficient (95%CI) | ICD10-AM, n | Comment |
|---|---|---|---|
| Rank-7 | 1.002, 1.135 | 464 | Dysphagia, fluid overload (oedema), tachypnoea, orhyponatraemia; osteomyelitis, “lower respiratory infection”, vascular/diabetic ulcer, trauma (chest, pelvis, thoracic spine, pathological fracture) endocrine tumour, low-risk poisons/drug overdose. |
| Rank-8 | 1.494, 1.633 | 268 | Acquired coagulopathy, acute delirium, lobar collapse, haematemesis or malaena; angiodysplasia, atherosclerosis, biliary obstruction, COPD ± infective exacerbation, colon cancer, diabetic ketoacidosis, SARS-COV2 infection, pancreatitis, pulmonary embolus, septic arthritis, spinal ischaemia, status epileptics, valvular heart disease. |
| Rank-9 | 1.780, 1.921 | 241 | Hypercalcaemia, ventricular tachycardia, pleural effusion, weight loss; congenital anaemias, breast cancer, prostate cancer, trauma (femoral, hip fracture, pneumothorax), high-risk poisons/drug overdose, rhabdomyolysis. |
| Rank-10 | 1.955, 2.096 | 95 | Arterial thromboembolism. cardiac failure, cholangitis, melanoma, non-traumatic brain injury, gasrtic or bowel obstruction, cutaneous pressure-injury, urological cancer. |
| Rank-11 | 2.111, 2.272 | 181 | Acute kidney injury, reduced conscious state; acute coronary syndrome, bacterial pneumonia, cardiomyopathy, cerebral abscess, carotid occlusion without stroke, dementia, hyperosmolar syndomes, myelodysplastic disorder, cancer (gastric, ovarian, uterine, oral), cervical spine trauma. |
| Rank-12 | 2.520, 2.735 | 256 | Chronic liver disease without cirrhosis, alcoholic hepatitis, empyema, Parkinsons, severe degenerative neurological disease, upper gastrointestinal perforation, pulmonary hypertension, cancer (throat or oesophagus) |
| Rank-13 | 2.806, 2.996 | 244 | Hypernatraemia, metabolic acidosis, lymphoma, severe burn injury, endocarditis, viral pneumonia, fungal pneumonia or sepsis, acute peritonitis, meningitis, interstitial lung disease, ischaemic stroke, cancer (of bone, lung, pancreas), subdural haematoma. |
| Rank-14 | 2.995, 3.201 | 212 | Acute respiratory failure, bacterial septicaemia, non-infectious encephalitis, myeloma, severe traumatic brain injury. |
| Rank-15 | 3.257, 3.481 | 51 | Acute hepatic failure, hypovolaemic shock, aspiration pneumonitis, chronic liver disease with cirrhosis, chronic kidney disease, leukaemia, mesenteric vascular disease, mesothelioma, cerebral cancer, encephalitis |

| Variable | ß coefficient (95%CI) | ICD10-AM, n | Comment |
|---|---|---|---|
| Rank-16 | 3.173, 3.449 | 45 | Metastatic cancer, necrotising fasciitis. |
| Rank-17 | 3.831, 4.194 | 18 | Aortic rupture, septic shock, chronic respiratory failure. |
| Rank-18 | 4.017, 4.312 | 21 | Haemorrhagic stroke, subarachnoid haemorrhage. |
| Rank-19 | 4.609, 5.055 | 11 | Cardiogenic shock, cardiorespiratory arrest. |
| Constant | -13.36, -12.20 | | |

**Table E3**

HOPE-7 model variables. Note: each record is permitted only one diagnosis Category derived from principal diagnosis; see main text for explanation. ß = model coefficient; ED = Emergency Department; CDG = Clinical Diagnosis Group[22], CFR= unadjusted case fatality rate.

**Example of HOPE-7 calculation.**

Three fictitious case examples of mortality risk prediction. Three 84 year-old males with the same three medical conditions (congestive cardiac failure, chronic kidney disease requiring ambulatory dialysis, and a large inguinal hernia) are admitted to a major hospital, but the reason for admission (principal diagnosis) differs for each, as indicated in Table E5.

| **Variable** | **ß (model coefficient)** | **Patient 1** | **Patient 2** | **Patient 3** |
|---|---|---|---|---|
| **Reason for admission** | | Cardiac failure | Ambulatory dialysis | Elective hernia repair |
| **Reason for admission, rank** | | 9 | 0* | 2 |
| **Age (years)** | | 84 | 84 | 84 |
| **Age, ß = coeff x (√age)** | 0.792 | 7.128 | 7.128 | 7.128 |
| **Male** | 0.105 | Yes | Yes | Yes |
| **Unplanned admission** | 0.721 | Yes | No | No |
| **Transfer to emergency department** | -0.482 | Yes | No | No |
| **Age-care resident** | 0.509 | Yes | No | No |
| **Transfer** | 0.852 | Yes | No | No |
| **Living alone** | 0.197 | Yes | Yes | No |
| **Diagnosis coefficient** | | 2.026 | 0 | -0.692 |
| **Model constant** | -12.776 | Yes | Yes | Yes |
| **Sum of coefficients = $\sum$ ß** | | -1.720 | -5.345 | -6.234 |
| **Predicted risk = $e^{ß}$** | | 0.179 | 0.005 | 0.003 |

**Table E5** Worked examples. *All low-risk CDG (rank-0) are given mortality risk of zero.

All cases are based on fictitious data and not derived from actual patients.

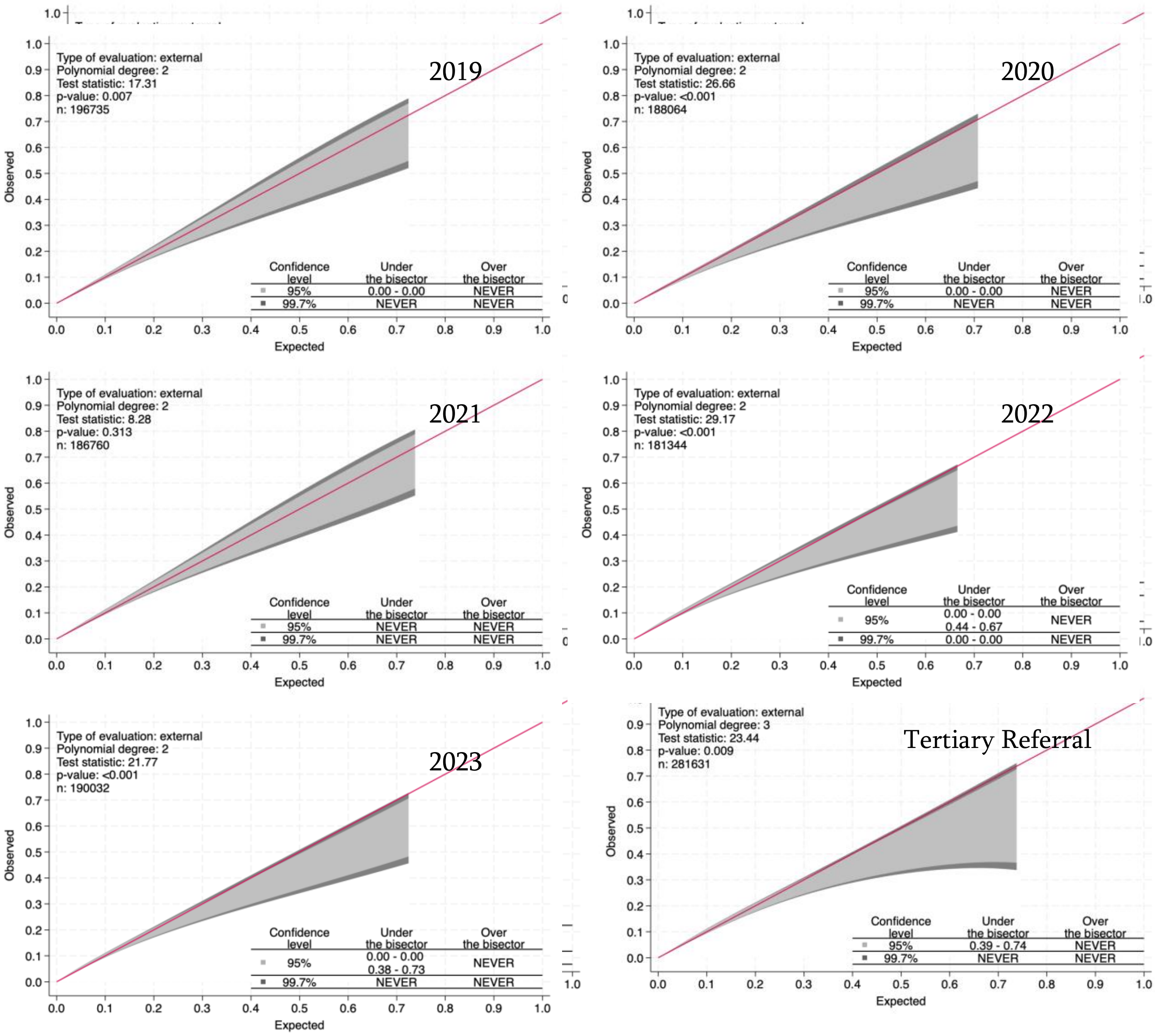


**Figure E2**

Calibration plots for HOPE-7 model across hospital peer groups in validation dataset.

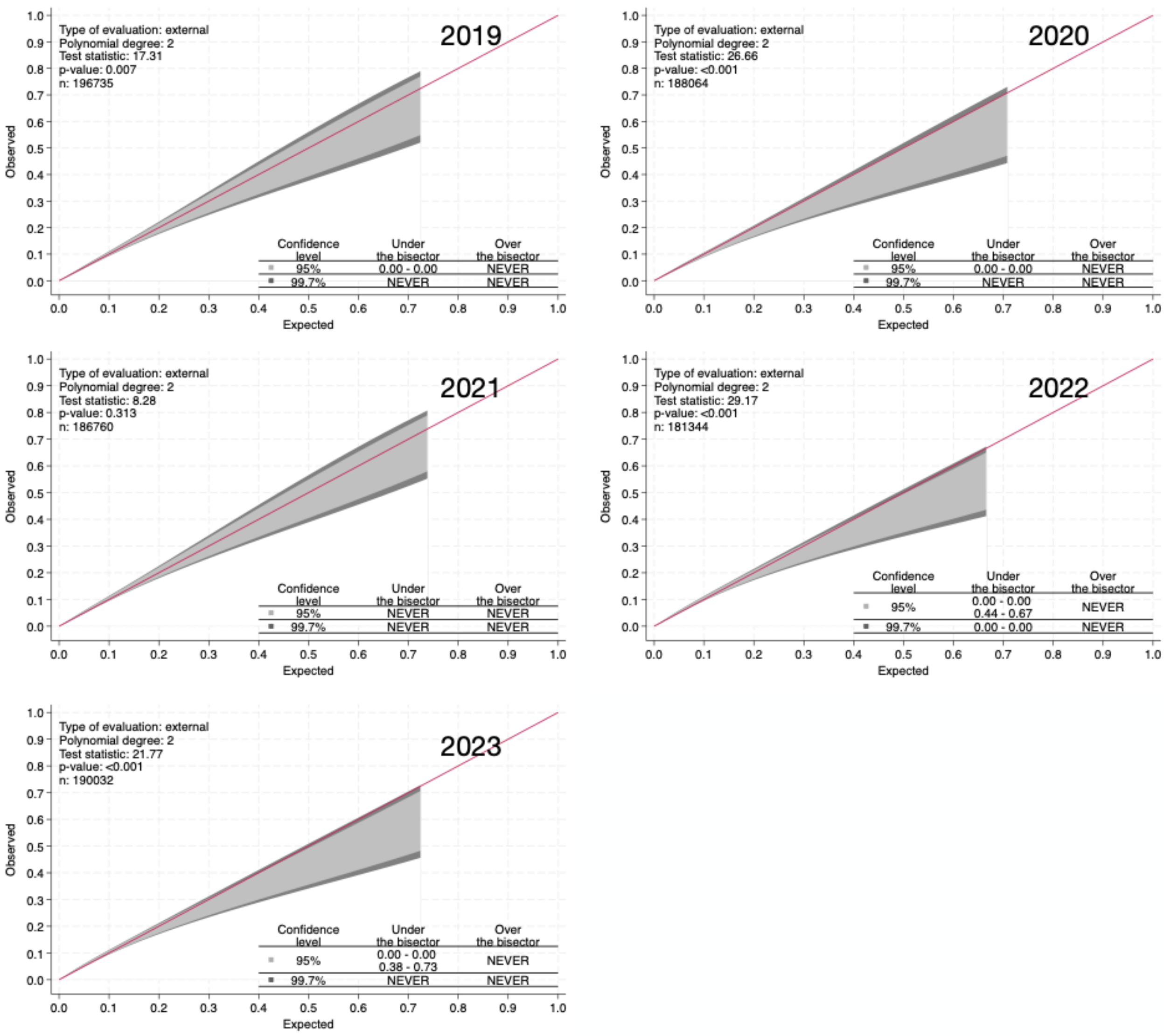


**Figure E3**

Calibration plots for HOPE-7 model across fiscal years (ending 30th June), validation dataset.

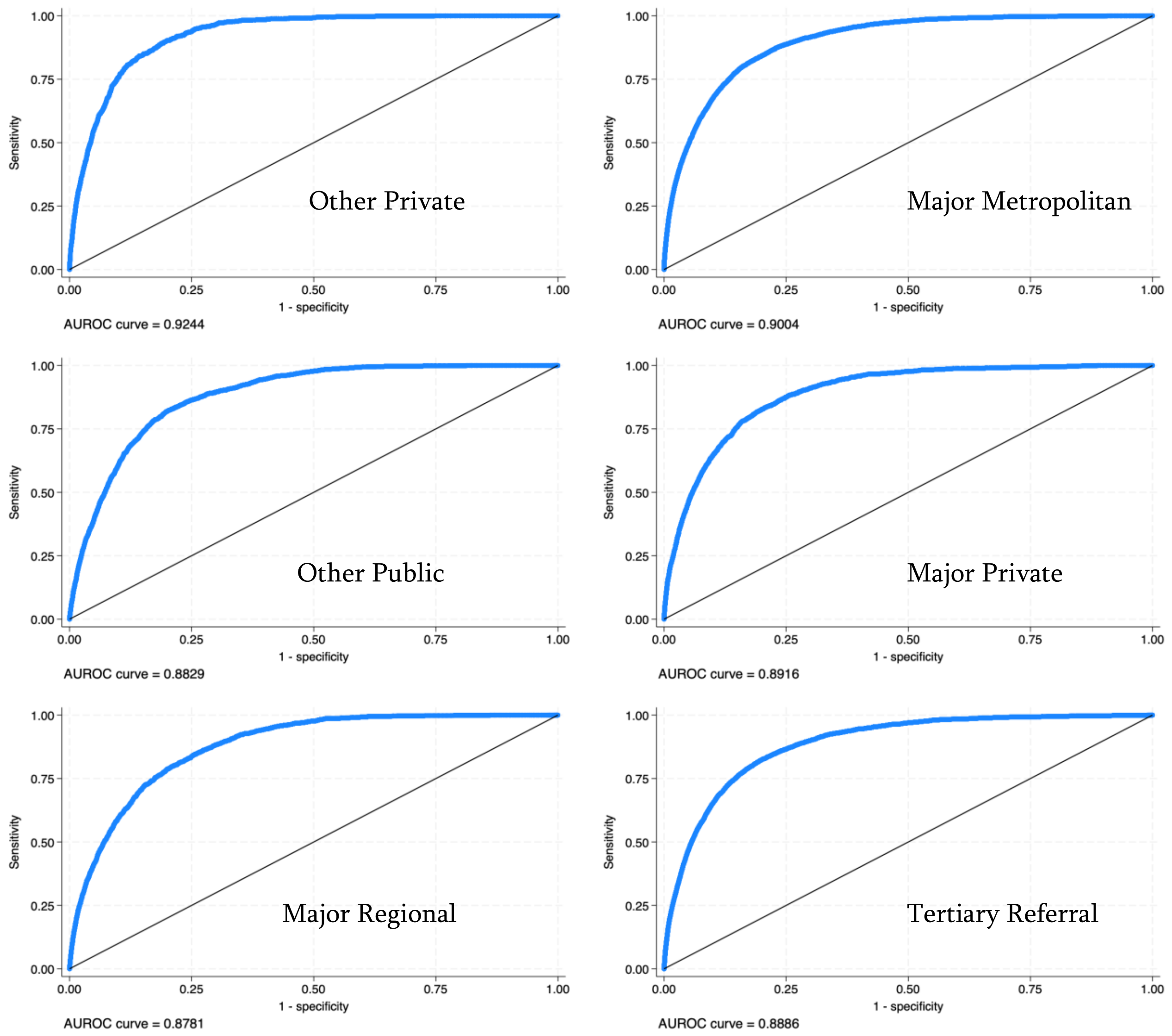


**Figure E4.**

Receiver-operator characteristic plots for HOPE-7 across hospital peer groups in validation dataset.

See Table 3, main text.

| **Fiscal quarter** | Hospitals, n | Hospital with no deaths, n | Separations n | Hospital deaths, n | $H_{10}$ | P-value | Dispersion value, φ | Dispersion RE SD, $\boldsymbol{\tau}$ |
|---|---|---|---|---|---|---|---|---|
| **2019Q1** | 277 | 139 | 649039 | 3908 | 18.9 | 0.015 | 3.6 | 0.22 |
| **2019Q2** | 279 | 146 | 662614 | 3687 | 13.3 | 0.102 | 4.5 | 0.25 |
| **2019Q3** | 281 | 143 | 639554 | 3454 | 14.9 | 0.061 | 4.4 | 0.26 |
| **2019Q4** | 282 | 149 | 660641 | 3840 | 8.3 | 0.409 | 4.0 | 0.24 |
| **2020Q1** | 279 | 141 | 672517 | 4025 | 12.4 | 0.133 | 3.5 | 0.23 |
| **2020Q2** | 278 | 144 | 672062 | 3720 | 13.2 | 0.107 | 3.4 | 0.20 |
| **2020Q3** | 278 | 147 | 637410 | 3506 | 18.9 | 0.016 | 3.3 | 0.21 |
| **2020Q4** | 272 | 140 | 530245 | 3369 | 11.1 | 0.194 | 3.8 | 0.24 |
| **2021Q1** | 283 | 144 | 569095 | 3868 | 18.4 | 0.019 | 4.8 | 0.28 |
| **2021Q2** | 282 | 145 | 666008 | 3359 | 13.4 | 0.100 | 4.3 | 0.23 |
| **2021Q3** | 279 | 146 | 656813 | 3396 | 16.7 | 0.034 | 3.8 | 0.22 |
| **2021Q4** | 271 | 134 | 676294 | 3770 | 17.4 | 0.026 | 4.4 | 0.24 |
| **2022Q1** | 278 | 145 | 701220 | 3920 | 15.1 | 0.056 | 4.7 | 0.24 |
| **2022Q2** | 273 | 141 | 623332 | 3912 | 10.1 | 0.255 | 5.1 | 0.25 |
| **2022Q3** | 274 | 142 | 586622 | 4028 | 11.0 | 0.203 | 5.0 | 0.25 |
| **2022Q4** | 271 | 143 | 656237 | 4182 | 18.3 | 0.019 | 4.3 | 0.23 |
| **2023Q1** | 275 | 139 | 668632 | 4487 | 35.8 | 0.000 | 4.5 | 0.24 |
| **2023Q2** | 269 | 136 | 679228 | 4077 | 9.5 | 0.305 | 5.8 | 0.26 |
| **2023Q3** | 269 | 140 | 670628 | 3909 | 6.6 | 0.583 | 4.4 | 0.23 |
| **2023Q4** | 265 | 136 | 691252 | 4213 | 14.4 | 0.073 | 4.4 | 0.23 |

**Table E5** Activity, calibration and dispersion metrics for each fiscal quarter derived from the validation cohort. $H_{10}$= Hosmer-Lemeshow chi-squared statistic for declines of equal size; P-value = probability of $H_{10}$ ; see main text for explanation and Figure E2, E3, E4.
